# Machine Learning for Predicting Therapeutic Outcomes in Acute Myeloid Leukemia Patients

**DOI:** 10.1101/2024.02.29.24303536

**Authors:** Nestoras Karathanasis, Panayiota Papasavva, Anastasis Oulas, George M Spyrou

**Affiliations:** Bioinformatics Department, The Cyprus Institute of Neurology & Genetics, 6 Iroon Avenue, 2371 Ayios Dometios, Nicosia, Cyprus; Molecular Genetics Thalassemia Department, The Cyprus Institute of Neurology & Genetics, 6 Iroon Avenue, 2371 Ayios Dometios, Nicosia, Cyprus

**Keywords:** Acute Myeloid Leukemia, Drugs response, Machine Learning

## Abstract

**Background and Objective:** The standard of care in *Acute Myeloid Leukemia* patients has remained essentially unchanged for nearly 40 years. Due to the complicated mutational patterns within and between individual patients and a lack of targeted agents for most mutational events, implementing individualized treatment for AML has proven difficult. We reanalysed the *BeatAML dataset* employing *Machine Learning algorithms*. The BeatAML project entails patients extensively characterized at the molecular and clinical levels and linked to drug sensitivity outputs. Our approach capitalizes on the molecular and clinical data provided by the *BeatAML dataset* to predict the *ex vivo* drug sensitivity for the 122 drugs evaluated by the project.

**Methods:** We utilized ElasticNet, which produces fully interpretable models, in combination with a two-step training protocol that allowed us to narrow down computations. We automated the genes’ filtering step by employing two metrics, and we evaluated all possible data combinations to identify the best training configuration settings per drug.

**Results:** We report a Pearson correlation across all drugs of 0.36 when clinical and RNA sequencing data were combined, with the best-performing models reaching a Pearson correlation of 0.67. When we trained using the datasets in isolation, we noted that RNA Sequencing data (Pearson: 0.36) attained three times the predictive power of whole exome sequencing data (Pearson: 0.11), with clinical data falling somewhere in between (Pearson 0.26). Lastly, we present a paradigm of clinical significance. We used our models’ prediction as a *health management score* to rank an individual’s expected response to treatment. We identified 78 patients out of 89 (88%) that the proposed drug was more potent than the administered one based on their *ex vivo* drug sensitivity data.

**Conclusions:** In conclusion, our reanalysis of the BeatAML dataset using Machine Learning algorithms demonstrates the potential for individualized treatment prediction in Acute Myeloid Leukemia patients, addressing the longstanding challenge of treatment personalization in this disease. By leveraging molecular and clinical data, our approach yields promising correlations between predicted drug sensitivity and actual responses, highlighting a significant step forward in improving therapeutic outcomes for AML patients.

**Highlights:**

- Machine learning can predict response to treatment in Acute Myeloid Leukemia patients.
- RNA sequencing data are more informative than whole exome sequencing and clinical data in predicting drug response in Acute Myeloid Leukemia patients.
- Drug response predictions could be used as a health management score to rank the individual’s expected response to treatment.
- We identified a more potent drug than the administered one for 88% (78 out of 89) of the patients examined.

## 1. Introduction

*Acute Myeloid Leukemia (AML)* is the most prevalent type of acute leukemia in adults. Over 20,000 cases are diagnosed yearly in the United States, leading to more than 10,000 deaths. Extensive chromosomal translocations and/or mutations in genes involved in hematopoietic proliferation and differentiation, lead to poorly differentiated myeloid cells with a survival advantage (leukemic blasts). AML is a highly heterogeneous disease with diverse outcomes; patients are classified as having favourable, moderate, or high-risk disease based on their cytogenetic and molecular profile, with prognostic implications. The discovery of recurring genetic abnormalities, such as the FLT3-ITD, NMP1 and CEBPA mutations, has refined individual prognosis and improved therapeutic strategies, particularly in determining the role of allogeneic stem cell transplantation (AlloSCT). Even so, induction treatment for fit patients still entails using an anthracycline + cytarabine backbone followed by consolidation with further chemotherapy or AlloSCT for those with high-risk disease. Elderly patients receive either supportive care or low-intensity regimens, such as low-dose cytarabine or hypomethylating agents (HMAs) alone or combined with Venetoclax, with poor outcomes[1]. Due to the complicated mutational patterns within and between individual patients and a lack of targeted agents for most mutational events, implementing individualized treatments for AML has proven difficult[2].

The *BeatAML project* connected the *ex vivo* drug sensitivity to patients’ molecular and clinical profiles[2]. *Every patient was extensively characterized at the molecular and clinical level and linked to drug sensitivity outputs*. Specifically, BeatAML characterized 672 specimens from 562 patients using whole-exome sequencing, RNA sequencing, detailed clinical data, and *ex vivo* drug sensitivity analyses. The *ex vivo* drug sensitivity analysis was performed across 122 drugs, and the Area Under the dose-response Curve (AUC) metric was calculated. Furthermore, patients were chosen to represent a wide range of categories, encompassing demographics (sex, age groups, and ethnicities), various karyotypic and molecular AML subtypes, as well as different clinical scenarios including cases of de novo AML, secondary AML, various types of preceding hematologic malignancies, and different types and stages of treatment. The BeatAML data are available in dbGap. Because of this multimodal characterization of its participants, compared to previous efforts, including The Cancer Genome Atlas (TCGA), and the Therapeutically Applicable Research to Generate Effective Treatments (TARGET), *BeatAML offers an original opportunity to create better patient stratification approaches*.

Nowadays, several efforts exist in employing machine learning to predict drug responses in AML using omics data. Lee et al. demonstrated a promising approach to identify robust molecular markers for targeted treatment of AML by introducing data from 30 AML patients, including genome-wide gene expression profiles and in vitro sensitivity to 160 chemotherapy drugs, a computational method to identify reliable gene expression markers for drug sensitivity by incorporating multi-omics prior information relevant to each gene’s potential to drive cancer[3]. Gerdes et al.[4] introduced Drug Ranking Using ML (DRUML) to generate organized lists of over 400 drugs based on their effectiveness in inhibiting cancer cell proliferation using omics data. To enhance the reliability of predictions while reducing unwanted variations, DRUML adopts internally normalized distance metrics of drug responses as features for creating machine learning models instead of relying on individual characteristics. The training data for DRUML comprises proteomics and phosphoproteomics data from 48 different cell lines, and they assessed its performance using data from 53 cellular models obtained from 12 independent research facilities. Trac et al.[5] developed MDREAM (Monotherapy Drug Response prediction for AML) using integrated omics data. MDREAM underwent training and initial validation utilizing the BeatAML cohort. The prediction process involves 122 ensemble models, each corresponding to a specific drug. The authors trained each model using Support Vector Machines and used RNA sequencing data as the input for the model. Furthermore, they selected features manually, including AML subtype-specific genes, pathway activation score, drug-target genes, mutated genes, and other AML-relevant genes. Additionally, they introduced a confidence score for each patient’s prediction to measure the prediction uncertainty for individual drugs, providing valuable guidance for practical clinical applications.

In this work, we re-analysed the BeatAML[2] dataset employing generalized linear models (*ElasticNet*[6]) to predict the area under the drug response curve, AUC. *ElasticNet* is a parametric method that fits generalized linear and similar models via penalized maximum likelihood[6]. The method has been used extensively in predicting drug sensitivity[7] and overall survival[8]. It is one of the most interpretable ML methods[9], performs an internal feature selection by removing highly correlated features and has a fast running time. The novelty of our analysis lies in the following characteristics. 1) We used multiple features and datatypes (Clinical, RNA sequencing, Whole Exome sequencing) alone or in combinations in order to train and test our models. 2) We implemented an automated data-driven approach for selecting features from the RNA sequencing data, using two different filtering metrics, and from the Whole Exome sequencing data, using one filtering metric. 3) We utilized a two-step training approach to narrow down computations and identify the best configuration settings per drug. 4) We implemented two complementary testing scenarios. In the first one, the same data are available for all training and testing samples. In the second scenario, some datatypes are missing, and our models predicted the AUC from the available datatypes. 5) We demonstrated the clinical utility of our approach by ranking patients’ expected response to treatment through the simultaneous use of *molecular and clinical attributes*. We showed that alternative drugs as the prescribed ones, albeit more potent, were predicted through this ranking.

## 2. Materials and Methods

### 2.1 Data

We trained and tested our models using the following data types.

*Whole-exome sequencing* is available for 369 samples. We used the genetic variants that passed all the filtering steps employed by the authors of the initial publication[2]. We utilized the HGVS nomenclature standard information, available in the supplementary data of the original publication, Table S7-Variants for Analysis, column *hgvsc*. To select which mutation events to retain, we optimized a parameter named *quantile_dnaseq*. For more information about the whole exome sequencing data see supplementary data, section Data.

*RNA sequencing* is available for 328 samples. We transformed the Counts Per Million (CPM) expression values, already available in the BeatAML dataset, according to the equation: *CPM*_*transformed*_ = *log*_2_(*CPM* + 10^−6^). We identified the best subset of genes by optimizing a parameter named *quantile_rnaseq* employing two filtering approaches: the *mean expression* or the *variance* of each gene. We used cross-validation to identify the best combination of the filtering method and *quantile_rnaseq* value; see the section *Training & Testing protocol* below. For more information about the RNA sequencing data see supplementary data, section Data.

*Clinical data* are available for 409 samples. After consulting our hematology expert, we used *categorical* and *numerical* data. Categorical data included ethnicity, sex, FLT3-ITD, NPM1 mutation status, Karyotype and others. Numerical data included age, hemoglobin, white blood cell count, and others. The meaning of each variable can be found in *Table S24-Clinical data diction* of the initial publication[2]. For more information about the clinical data see supplementary data, section Data.

### 2.2 Variable to Predict

We used the *Area Under the dose-response Curve (AUC)* as the response variable, that is the variable we trained our models to predict. Both AUC and IC50 values are available for *410 samples* and *122 drugs*. The number of available samples per drug ranges from 80 to 399. AUC captures potency information (EC50, IC50) and drug efficacy (Amax) by a single measure. Previous analyses showed AUC to be a robust metric for comparing a single drug across cell lines and a better standard of cell line selectivity compared to IC50[10].

### 2.3 Training and Testing Protocol

We utilized ElasticNet to construct our models using its implementation in R’s glmnet package. We minimized the *Mean Square Error (MSE)* between the predicted (*auc_hat*) and the true AUC value in order to select a) the best combination of configurations and hyperparameters during training and b) the best-performing model.

In the beatAML dataset, different numbers of samples are available per drug and datatype. For example, the “17-AAG (Tanespimycin)” drug has been measured in 344 samples. Clinical, whole exome sequencing, and RNA sequencing data are available for 344, 308, and 284 samples, respectively. All data types are available for 251 samples.

We set up our training-testing protocol to take into account all samples for which an AUC measurement is available in the data. As a result, training and testing can consider different input data and produce an AUC prediction (*auc_hat*) for every sample. We performed the following steps to train and test our models separately for every drug.

1. *Split the data* into ten folds. All folds equally represent the initial distributions of the available data. For example, for the “17-AAG (Tanespimycin)” drug, 80% (284 / 344), 89% (308/344) and 100% of the samples had RNA sequencing, whole exome sequencing and clinical data available, respectively. We maintained similar percentages in each fold. We selected nine folds (90% of the data) for *training* and one-fold (10% of the data), namely the *outer cross-validation fold* for *testing*. This data splitting aimed at a *nested cross-validation* protocol to estimate the performance of our analysis.
2. *Training*. We used only the training data in the following steps.
  2.1. *Two-step training*. We trained our models in a two-step manner. In the first step, we trained three models, one for each data type (*clinical, rnaseq, variants*). In this step, we explored all possible combinations of hyperparameters to optimize; see section 2.2. We selected three models, one per datatype, that produced the lowest cross-validated Mean Square Error *(cvMSE)*. In the second step, we stabilized all the hyperparameters identified in the first step except glmnet’s hyperparameters (*alpha* and *lambda*), and we trained four more models by integrating the available datatypes in all possible combinations. In this step, we re-optimized the glmnet’s hyperparameters. As the sample sizes between datasets differ, we used the maximum overlap every time.
  2.2. *Hyperparameter optimization. Glmnet* has a set of parameters called hyperparameters that the user needs to optimize: *alpha* and *lambda*. In this set, we included *quantile_rnaseq, quantile_dnaseq*, and *tumor_only* (defined above - sections RNA sequencing and whole-exome sequencing). All hyperparameters were optimized using a 10-fold cross-validation[9]. In the first training step, we optimized *quantile_rnaseq, quantile_dnaseq, tumor_only*, and glmnet’s *alpha* and *lambda*, and in the second training step, we kept *quantile_rnaseq, quantile_dnaseq, tumor_only* constant and we reoptimized only glmnet’s hyperparameters.
  After training, we had seven models for each drug: three that employed only one data modality and four that employed their combinations.
3. *Testing*. After training, we calculated the performance of our models in the *outer cross-validation fold* using Mean Square Error (*nestedcvMSE*), Pearson, and Spearman correlations between the true AUC and the *auc_hat* prediction. We performed the testing using two complementary ideas, namely *same_input* and *different_input*. We evaluated the seven models separately in the *same_input* case, where the input is the same in the training and testing phase. In the *different_input* case, we evaluated the performance of the models considering *all samples* in the *outer cross-validation* fold. Each sample could have one or all data types available in this case. We predicted *auc_hat* using the best out of the available models. We identified the best model using their *cvMSE* performance. For example, if a sample has all datatypes, clinical, RNAseq, whole exome, available, we predicted its *auc_hat* using the best out of the seven models, *clinical, rnaseq, variants, clinical+rnaseq, clinical+variants, clinical+rnaseq+variants*. If a sample had only clinical and RNAseq data available, we predicted its *auc_hat* using the best model out of the three available, *clinical, rnaseq, clinical+rnaseq*.
4. We performed steps 2 and 3, 10 times moving the testing fold across the several data splits, resulting in a ten-fold nested cross-validation scheme that has been proven to provide robust and unbiased performance estimates[11].

## 3. Results

### 3.1 Performance

#### 3.1.1 Overfitting Evaluation

First, we evaluated if our models overfitted the training set by comparing the models’ MSE during cross-validation (*cvMSE*) with the MSE in the outer holds of the nested cross-validation (*nestedcvMSE*). Specifically, *cvMSE* is the cross-validation MSE achieved by the model during training, and *nestedcvMSE* is the MSE achieved by the model in the outer-*unseen* set during the nested cross-validation. We calculated the mean cvMSE (*mean_cvMSE*) and the mean nestedcvMSE (*mean_nestedcvMSE*) across the ten nested cross-validation folds per datatype and drug and compared the two metrics. The median of the *mean_nestedcvMSE*’s distribution is slightly higher than the median of the *mean_cvMSE*’s distributions, which is expected for all datatypes and datatype combinations, Figure 1. Furthermore, the two metrics are highly correlated, with their Pearson correlations being higher than 0.96 in all cases, Supplementary Figure 1.

**Figure 1.**
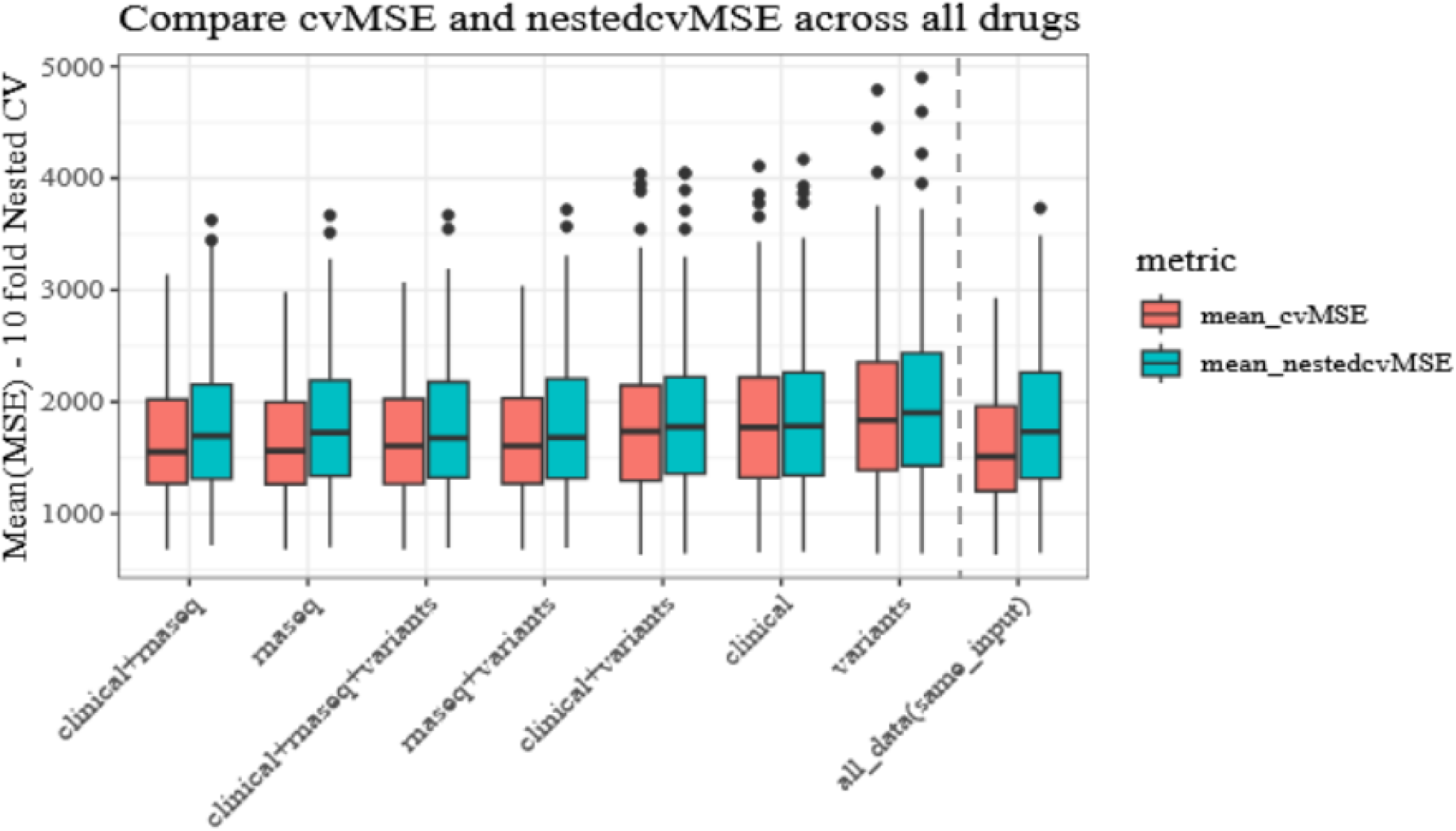
Overfitting Evaluation. We evaluated overfitting by comparing mean_cvMSE and mean_nestedcvMSE across all drugs, datatypes and datatype combinations. In all cases, our models show a good generalization performance.

#### 3.1.2. Same input setup

In the *same_input* setup idea, we emulated the case where the same datatype or datatype combinations are available for all samples employed in training and testing. We wanted to evaluate the question of how well a model performs if only one type of data was available, *clinical, rnaseq, variants* versus their combinations, *clinical+rnaseq, clinical+variants, rnaseq+variants, clinical+rnaseq+variants*. In all cases, we used the maximum shared number of available samples. Per drug, we trained seven models, predicted the *auc_hat* of the outer cross-validation test set, and calculated the Pearson and Spearman correlation between *auc_hat* and the true AUC. We performed this process ten times and calculated the mean Pearson (*mean_pearson*) and mean Spearman (*mean_spearman*) across these runs.

*Variants* showed the worst performance with a median *mean_pearson* of 0.112 across all drugs, Figure 2, with all the observed differences to be statistically significant (Wilcoxon test, Supplementary Table 1). *Clinical+variants* and *clinical* came right after with a median *mean_pearson* performance of 0.258 and 0.26, respectively. The difference between them is not statistically significant (Wilcoxon test, supplementary Table 1). Adding the RNAseq data in the mix boosted the performance above 0.3, Figure 2. *Rnaseq+variants* and *clinical+rnaseq+variants* have a median *mean_pearson* of 0.328 and 0.341, respectively. RNAseq (*rnaseq*) data alone come second from the top with a median *mean_pearson* of 0.357, and the best is the *clinical+rnaseq* with a median *mean_pearson* of 0.360, Figure 2. The differences observed within the models using RNAseq data were not statistically significant. However, all the observed differences between models with and without RNAseq data were statistically significantly different, supplementary Table 1.

**Figure 2.**
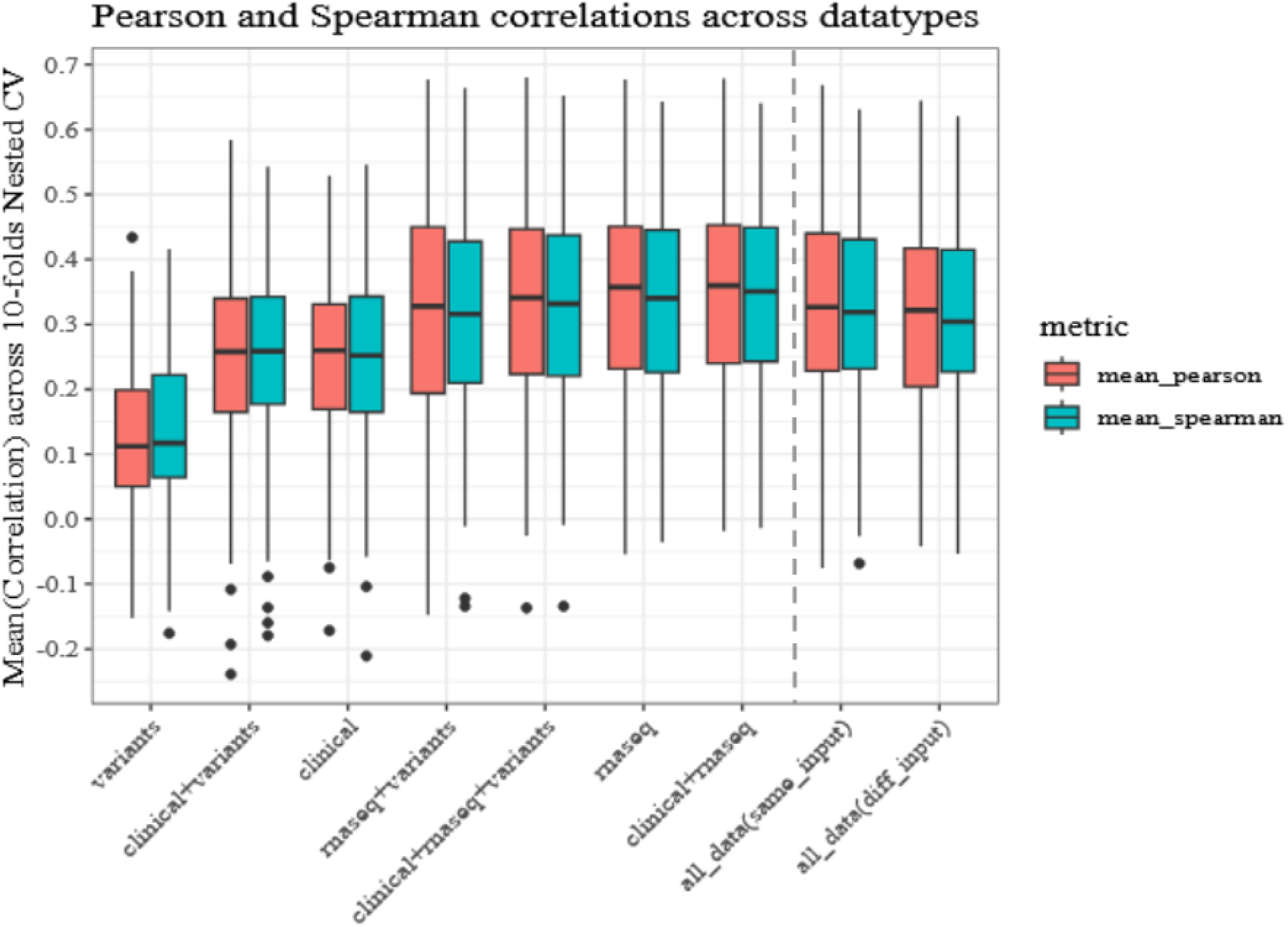
Pearson and Spearman correlation performance. Mean_pearson and mean_spearman across the various datatypes and datatype combinations for all drugs. X axis is sorted from the lowest to the highest performing datatype or datatype combinations except for all_data(same_input) and all_data(diff_input), on the right of the dashed grey vertical line.

Furthermore, instead of using the same datatype or datatype combination across all drugs, we selected the best datatype or datatype combination per drug using the *cvMSE* and evaluated if this approach would increase the performance. Similarly, we calculated the performance utilizing the *mean_nestedcvMSE, mean_pearson* and *mean_spearman* metrics. We achieved a median *mean_nestedcvMSE* equal to 1733.6046, Figure 1, a median *mean_pearson* equal to 0.327, and a median *mean_spearman* of 0.319, Figure 2. Only the differences between *all_data(same_input)* and *variants, clinical+variants*, and *clinical* are statistically significant. The performance of the selected models ranged from a median *mean_pearson* and *mean_spearman* of -0.075 and -0.0683 with the best to have median *mean_pearson* and *mean_spearman* correlations equal to 0.67 and 0.63, respectively, Figure 2, boxplots above “all_data(same_input)” label. We noted that based on the performance of the external sets, this approach came fifth using the *mean_nestedcvMSE* or the *mean_pearson* metrics and fourth using the *mean_spearman* without the observed differences being statistically significant, supplementary Table 1. However, based on the *cvMSE* metric we used to select the best models, this approach produced the lowest median *mean_cvMSE*, Figure 1. All drugs with their respective nested cross-validation Pearson correlations are available in Supplementary Figure 2.

#### 3.1.3. Different inputs setup

In the *different_input* setup idea, we evaluated the scenario where different types of data were available per sample, and we provided one prediction from the best available model. Specifically, different types of data are available per sample and drug. For example, clinical data are available for 79 samples in the case of Entrectinib but 398 in the case of Imatinib, as shown in Figure 3 and Supplementary Table 2. The more data types we merged, the fewer samples we had available. For example, in the case of A.674563 drug, 355, 319, 293, and 258 samples are available for *clinical, clinical+variants, clinical+rnaseq*, and *clinical+rnaseq+variants*, respectively. We observed similar patterns for the other drugs. Thus, to provide an *auc_hat* prediction for all samples in the outer cross-validation test set, we predicted *auc_hat* employing the best available model (see methods, section Training & Testing protocol). In this case, the median *mean_pearson* correlation is 0.322, and the median *mean_spearman* is 0.304, Figure 2, boxplot with x-axis label *all_data(diff_input)*. Only the observed differences between the *all_data(diff_input)* and *variants, clinical+variants* and *clinical* were statistically significantly different. For the per-drug Pearson correlation across the 10-fold nested cross-validation, see Supplementary Figure 3.

**Figure 3.**
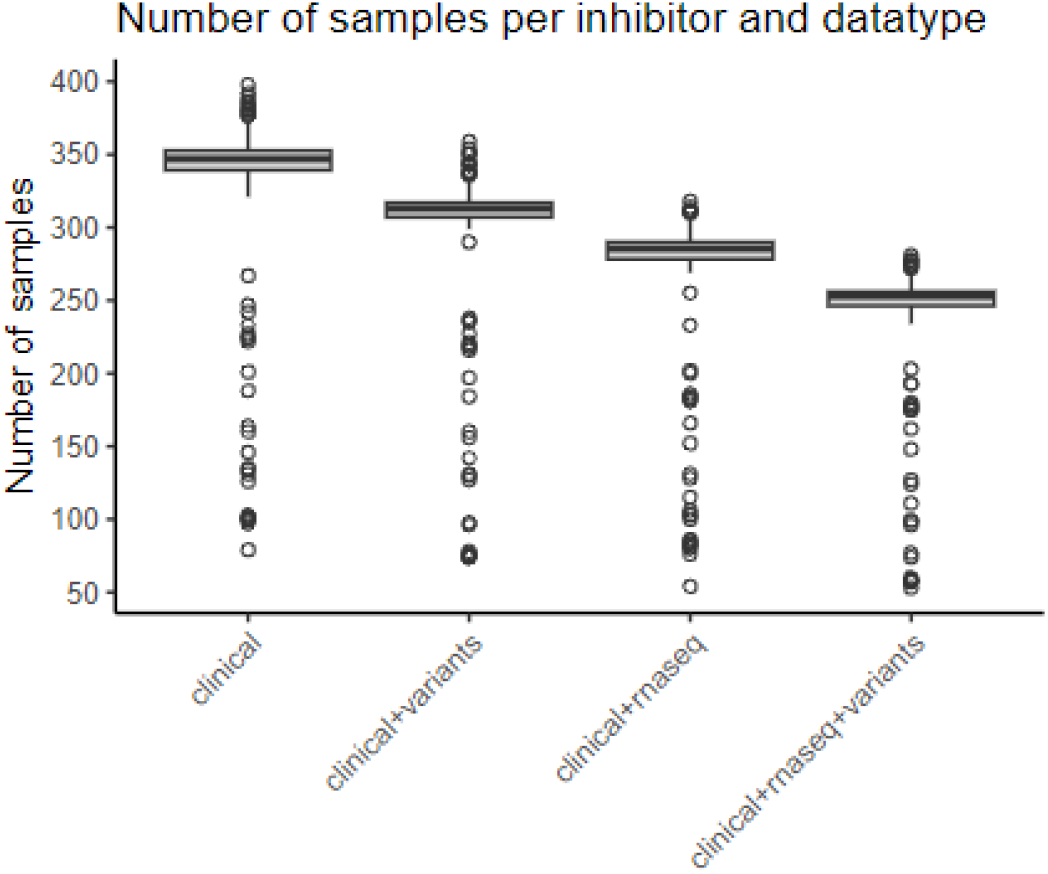
Diverse numbers of samples are available per drug and datatype.

### 3.2 Model interpretation

In this part, we interpreted our models concerning the datatypes and the respective configurations selected across drugs.

#### 3.2.1. Different datatypes or datatype combinations selected for each drug

In the case of the *same_input* setup, we allowed our algorithm to select the best datatype or datatype combination per drug. The *clinical+rnaseq* datatype combination was selected most of the times, 298 out of the 1220 runs (10-fold nested cross-validation * 122 drugs). *Rnaseq* was selected 268 times and the use of all datatypes, *clinical+rnaseq+variants* was selected 218 times. The *rnaseq+variants, clinical+variants, clinical*, and *variants* datatypes were selected in 176, 151, 86, and 23 runs, respectively, Figure 4. It is worth noting that *clinical* and *variants*, the datatypes currently employed in clinical practice, were selected the least times and showed the worst performance.

**Figure 4.**
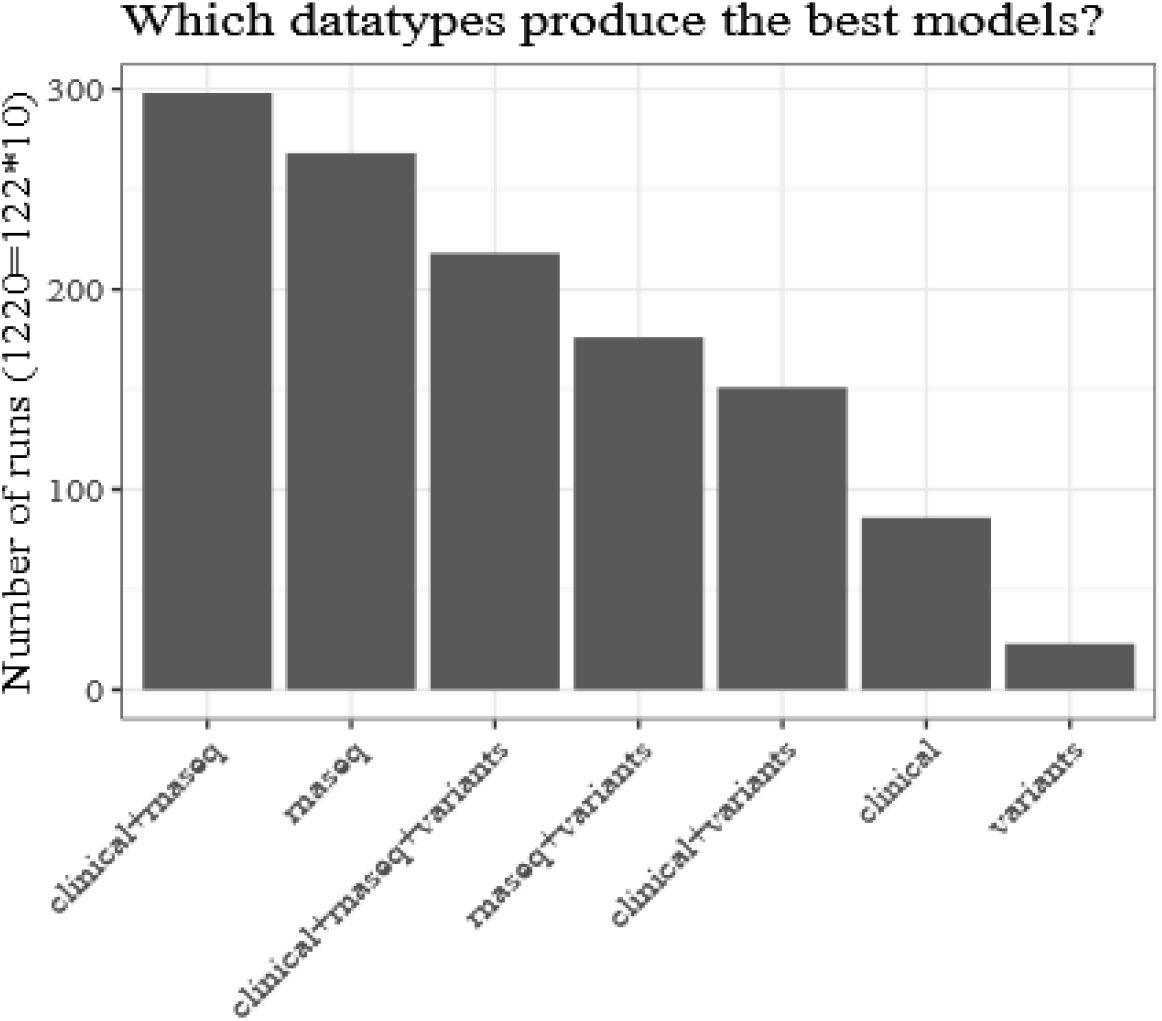
Datatypes selection summary. Number of times a datatype or datatype combination was selected by the best models during the nested cross-validation process across all drugs.

Furthermore, our algorithm selected different datatype combinations during the 10-fold nested cross-validation procedure for the same drug, Figure 5. For example, for the Afatinib-BIBW-2992 drug, our algorithm deviated a lot in the datatype selections. It selected *clinical+rnaseq* two times, *clinical+rnaseq+variants* two times, *clinical+variants* two times, *rnaseq+variants* one time and *variants* three times. On the other side of the spectrum, for the GDC-0879, our algorithm selected the *clinical* data all ten times. Our algorithm selected different datatypes across the 10-fold nested cross-validation for most drugs. We clustered the datatypes and the drugs, employing hierarchical clustering with Euclidean distance and the complete clustering algorithm, based on the frequency that each datatype was selected for model building for each drug, see Figure 5. Datatypes produced four main clusters when we cut at height equal to 36. The first cluster consisted of *clinical, variants* and *clinical+variants*, the second of *clinical+rnaseq*, the third of *clinical+rnaseq+variants* and *rnaseq+variants* and the fourth of *rnaseq*. Drugs generated five main clusters when we cut at height equal to 12. Starting from the top of figure 5, the first group contained drugs that mainly used *clinical* data. Drugs in the second group were more likely to use *clinical+variants*, whereas drugs in the third, fourth, and fifth groups were more likely to use *clinical+rnaseq, rnaseq*, and *clinical+rnaseq+variants and rnaseq+variants*, respectively, Figure 5.

**Figure 5.**
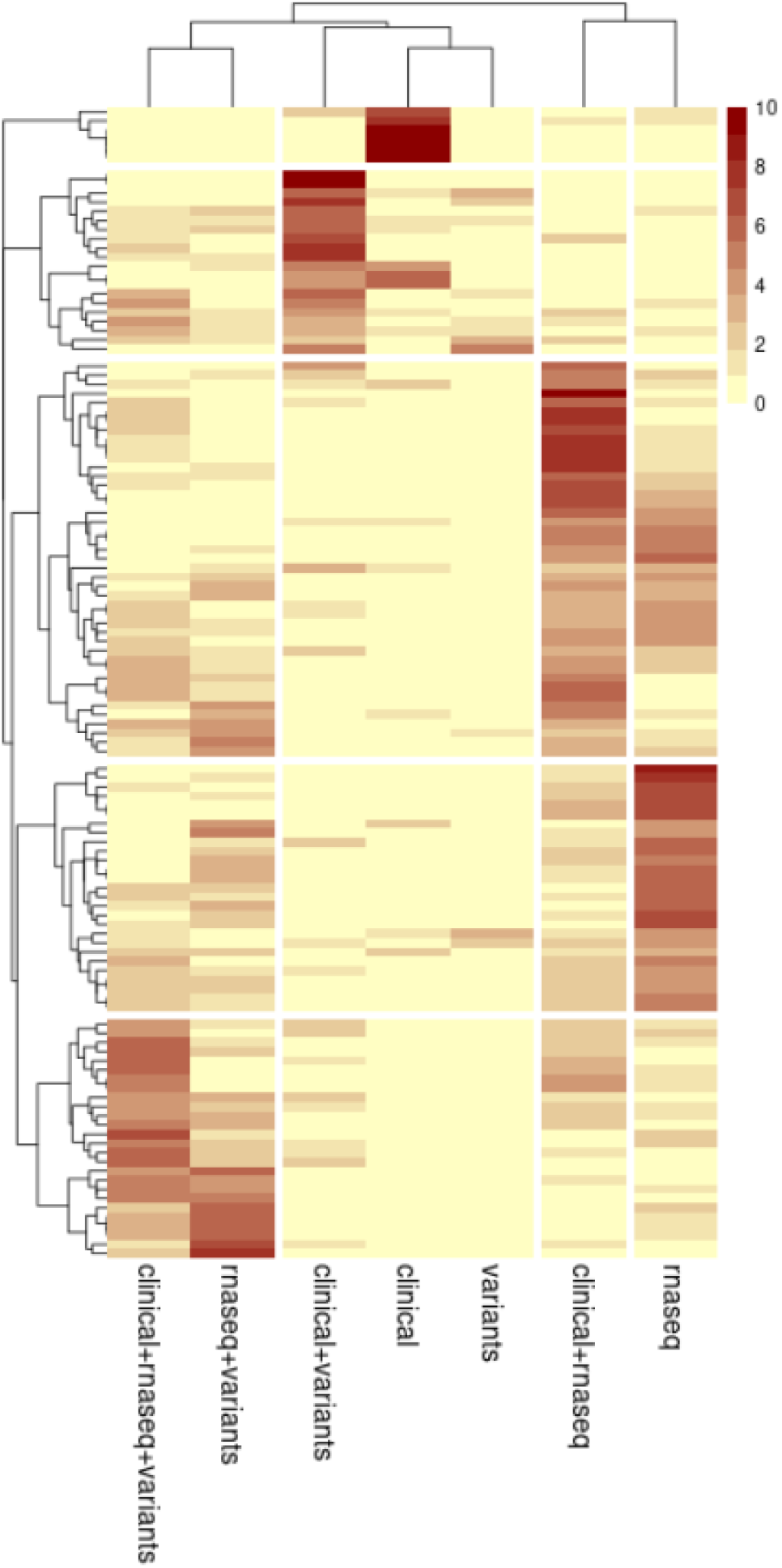
Datatype’s selection. The number of times a datatype combination was selected for a drug for model building during the ten-fold nested cross-validation. Yellow corresponds to zero times and dark red to ten times. Datatypes produced four main clusters that were separated by white vertical lines. Drugs generated five main clusters that were separated by white horizontal lines.

Furthermore, we mapped the drugs to their corresponding *drug families* as provided in the initial publication[2] and evaluated if any patterns emerged. We noted that seventy-one drugs belong to one family each, and the remaining fifty-one belong to several families. The number of families in some cases reached seven, eight, or nine, Supplementary Figure 4. Also, the number of drugs per family differs. Ten families have one drug, four have two, and some have up to 25 drugs, Supplementary Figure 5. To normalize for these differences, we calculated the fraction of the times a datatype was selected by a specific family during the ten-fold nested cross-validation. We did not observe any patterns that could be due to the drug family and not the individual drug present in the data, Supplementary Figure 6. For example, the GSK3 family showed a distinctive pattern where only clinical data were selected across all runs. However, only one drug corresponds to the GSK3 family. Similarly, families with one or few drug members achieved higher maximum fractions than families with many. The Spearman correlation between the number of drug family members and the maximum fraction achieved is -0.63, which is statistically significant (p-value: 1.59e-05), Supplementary Figure 7. For a detailed analysis of the RNAseq and whole exome sequencing configurations selected for each drug see supplementary data, sections *Different RNASeq configurations selected for each drug* and *Different Whole Exome configurations selected for each drug*, respectively.

### 3.3. Clinical implications

Lastly, we evaluated the practical utility of models that predict the response to treatment. To this end, we utilized the AUC predictions as a *health management score* to rank an individual’s expected response to treatment. Based on this score we suggested alternative drugs that are expected to be more effective than the administered ones. To perform this analysis, we selected drugs that are present in the patients’ cumulative treatment regimen, for which the response measurements (AUC) are available for the same patients. Eighty-nine patients and seven drugs fulfilled these requirements, namely Crenolanib, Dasatinib, Imatinib, Lenalidomide, Midostaurin, Sorafenib and Sunitinib. We noted that each administered drug has a specific number of alternatives based on the availability of *ex vivo* drug sensitivity data.

For example, in the case of Sorafenib, 52 patients were eligible for this analysis, Supplementary Figure 15, and the number of alternative drugs for each patient ranged from eight to one hundred three, Supplementary Figure 16. The same applies to the other drugs. Then, for every patient, we suggested, as an alternative drug, the drug with the lowest *auc_hat* prediction. We calculated the difference between the *true AUC* of the recommended and the *true AUC* of the administered drug, Figure 6. For four drugs (Crenolanib, Imatinib, Lenalidomide and Sunitinib), all patients showed a lower true AUC on the suggested versus the administered drugs. For three drugs (Dasatinib, Midostaurin and Sorafenib), the majority of the patients showed the same trend. The respective AUC distributions of the recommended versus the administered drugs are in Supplementary Figure 17.

**Figure 6.**
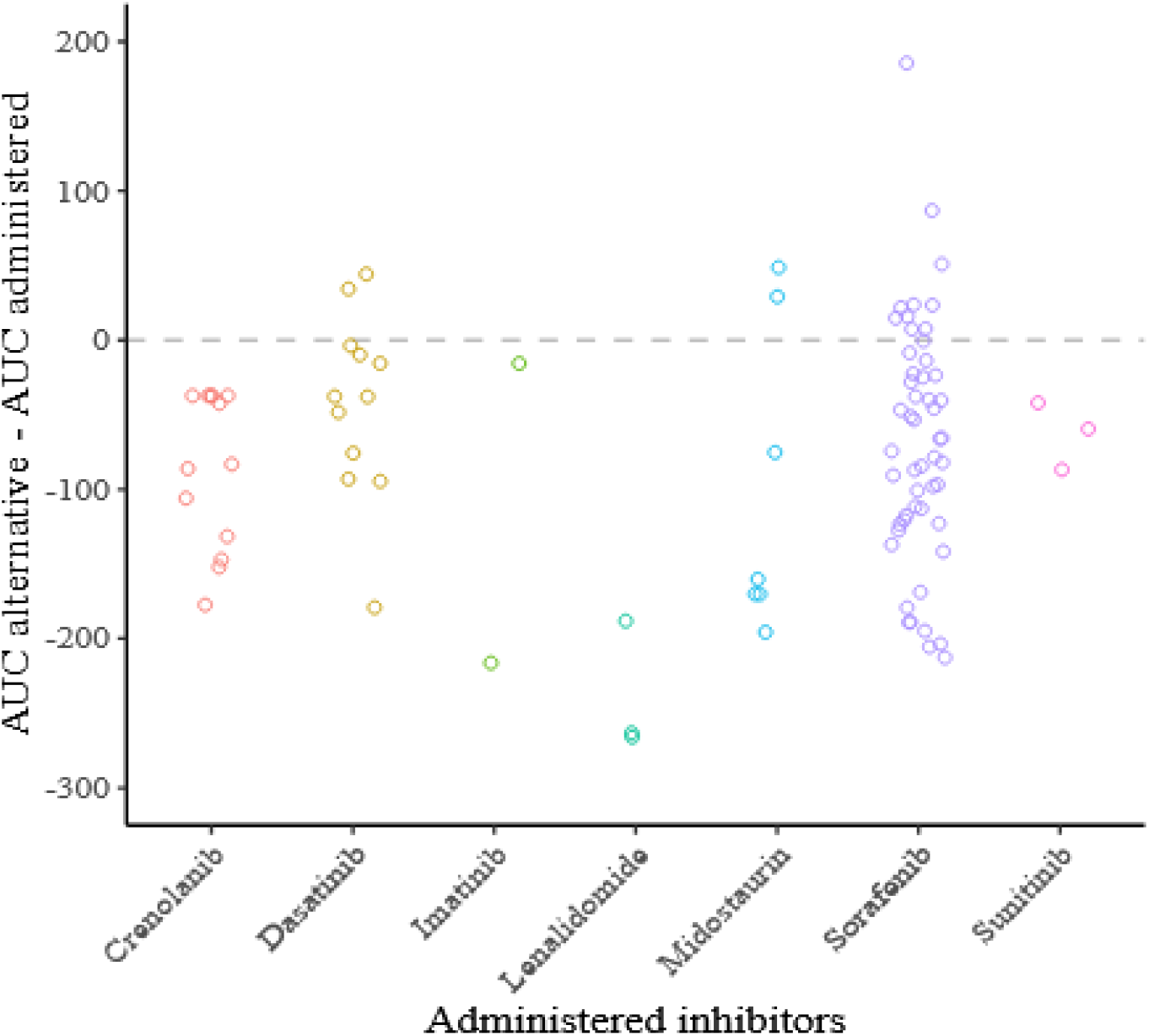
Clinical implications paradigm. The difference between the true AUC of the alternative drugs minus the true AUC of the administered drugs. Every point in the graph corresponds to a patient, 89 patients in total. For 78 patients, points below zero (grey reference line), the alternative drug has a lower AUC (more drastic) than the administered ones, x-axis.

## 4. Discussion

In this study, we reanalysed the beatAML dataset and generated machine-learning models that predict the expected response to treatment in the Area Under the drug response Curve (AUC) units. For model building, we utilized all available data types, namely clinical, RNA, and Whole Exome sequencing, in isolation and all possible combinations. Our analysis showed similar overall performance compared to previous efforts[5]. Specifically, Quang et al.[5] used a single hold-out approach to evaluate performance and reported a median correlation of 0.35. Even though they used information from the Whole Exome and RNA sequencing data, they utilized only the RNA sequencing as input in the training process. Our median correlation across a ten-fold nested cross-validation using only RNAseq is 0.357. When we used both RNAseq and clinical data, we achieved a median correlation of 0.360. In addition, Quang et al. reported an extensive feature extraction process by employing prior knowledge and a data-driven approach. In our analysis, we automated this part by optimizing two parameters: the *quantile_rnaseq*, which identified the best proportion of genes to retain based on the RNA sequencing data, and the *quantile_dnaseq*, which identified the best subset of mutation events to include based on their frequency in the Whole Exome sequencing. Importantly, ElasticNet produces fully interpretable models compared to Support Vector Machines[9]. Models’ interpretability is of critical importance in clinical applications.

Furthermore, we used two different scenarios in our analysis. In the *same_input* setup, we assumed all data were available for all samples. In the *different_input* setup, our implementation predicted AUC for all samples given the available data. Through the *same_input* scenario, we observed that the Whole Exome sequencing data showed the worst performance compared to RNAseq or clinical data alone. Interestingly, and as it has been shown in previous studies, RNA sequencing data perform better in drug response prediction tasks[7]. In this analysis, RNA sequencing data performed three times better than the Whole Exome data. By the *different_input* scenario, we estimated a more realistic performance in the clinical settings where some data might be missing for some patients. Our median correlation was 0.322, as it is a combination of predictions of models that use any type of data combinations.

In addition, we discuss a possible application of this type of analysis in a *clinical support decision system* setting. To highlight this idea, we analyzed 89 patients who were administered a drug for which alternative drugs are available in the data. By alternative drugs, we mean drugs whose *ex vivo* drug sensitivity (AUC) is available for the same patients. We predicted auc_hat using their molecular and clinical profile, and we employed this prediction as a *health management score* to rank an individual’s expected response to treatment. We identified 78 patients (88%) for whom the alternative drug was more drastic than the administered one based on their *ex vivo* drug sensitivity data.

Our implementation has several limitations and possible extensions. We focused our analysis on using ElasticNet. We did not evaluate the performance of other machine learning methods commonly employed in drug response prediction tasks, such as Random Forest, Support Vector Machines, Deep Learning, and others. For example, the poor performance of the Whole Exome sequencing data could be due to the selected method to build the models or the way we encoded that information. In the future, we will evaluate the performance of other methods and try different normalizations and encodings for the RNA and the Whole Exome sequencing data. About the clinical utility of these methods, our analysis assumed a monotherapy scenario where one drug is administered to a patient. However, in the clinical setting, a combination of drugs (for example, 7+3 (Cytarabine, Idarubicin)|MiDAC|AC220 (Ambit), etc.) is utilized in the different stages of the disease (e.g., Initial Acute Leukemia Diagnosis, Relapse, Post-Chemotherapy, Residual Disease, Unknown, Post-Transplant, Residual Relapse, Post-DLI). Future machine learning models could consider drug-to-drug interactions and suggest possible drug combinations that target dysregulated complementary disease pathways based on a patient’s molecular profile.

In conclusion, our study represents a significant step towards overcoming the longstanding challenges in Acute Myeloid Leukemia treatment by integrating advanced Machine Learning techniques with rich molecular and clinical datasets. The potential for personalised treatment strategies based on ex vivo drug sensitivity predictions open new avenues for improving therapeutic outcomes, and it marks a promising direction for future research and clinical applications in Acute Myeloid Leukemia.

## Data Availability

All data produced are available online at the supplementary material of the initial publication: https://www.nature.com/articles/s41586-018-0623-z,
under the section "Supplementary Tables" and can be found here: https://static-content.springer.com/esm/art%3A10.1038%2Fs41586-018-0623-z/MediaObjects/41586_2018_623_MOESM3_ESM.xlsx

## Abbreviations

(AML): Acute Myeloid Leukemia
(AlloSCT): Allogeneic Stem Cell Transplantation
(HMAs): hypomethylating agents
(AUC): Area Under the dose-response Curve
(TCGA): The Cancer Genome Atlas
(TARGET): Therapeutically Applicable Research to Generate Effective Treatments
(LASSO): regularized regression modelling
(DRUML): Drug Ranking Using ML
(CPM): Counts Per Million
(RPKM): Read Per Kilobase Million
(MSE): the Mean Square Error
(auc_hat): AUC prediction
(cvMSE): cross validated MSE
(nestedcvMSE): nested cross-validation
(mean_nestedcvMSE): mean nestedcvMSE
(mean_pearson): mean Pearson
(mean_spearman): mean Spearman

