## Supplementary material for "Machine Learning for Predicting Therapeutic Outcomes in Acute Myeloid Leukemia Patients"

### Data

Whole-exome sequencing is available for 369 samples. We used the genetic variants that passed all the filtering steps employed by the authors of the initial publication[2]. We utilized the HGVS nomenclature standard information**,** available in the supplementary data of the original publication, Table S7-Variants for Analysis, column hgvsc. We transformed this information using the "one-hot" encoding[9]. In initial experimentation, we observed that hgvsc includes 4912 distinct variants. The number of samples with a specific variant (frequency) range from 1 to 109, with most of the mutations (2435) appearing only in two samples. To select the mutation events to retain, we optimized a parameter named quantile_dnaseq. This parameter ranged from 0 to 1 and encoded the quantile of the frequency distribution that we used in our analysis. For example, when quantile_dnaseq = 0, we used all variants. When quantile_dnaseq = 1, none of the variants were used. When quantile_dnaseq = 0.5, we used the variants whose frequency was higher than the median of the frequency distribution. We set quantile_dnaseq to 0, 0.5, and 0.9. As an extra filtering step for variant selection, we used the tumor_only variable available in the supplementary data of the original publication. We employed a cross-validation procedure to optimize percentile_dnaseq in combination with tumor_only; see the section Training & Testing protocol below.

RNA sequencing is available for 328 samples. We transformed the Counts Per Million (CPM) expression values, already available in the BeatAML dataset, according to the equation: $\mathrm{CPM}_{\mathrm{transformed}} = \log_2(\mathrm{CPM} + 10^{-6})$. This choice serves two purposes. Firstly, the CPM expression of a gene is better comparable across samples versus the Read Per Kilobase Million (RPKM) normalization that is highly variable among samples[10]. Secondly, the use of $\log_2$ allows for the CPM expression values to be approximated with the Gaussian distribution versus the Negative Binomial distribution. We added $10^{-6}$ before calculating the $\log_2$ to avoid infinity values in our data.

Gene expression level is highly variable in the data. Selecting genes present in at least a specific fraction of samples or above an expression level is typical in differential expression analysis. Our preliminary analysis shows that such filtering would benefit our models. We identified the best subset of genes by optimizing a parameter named quantile_rnaseq employing two filtering approaches: the mean expression or the variance of each gene. quantile_rnaseq ranged from 0 to 1 and corresponded to the quantile of the distribution of the mean expression or the variance of the genes to retain. For example, when quantile_rnaseq = 0, we kept all genes. When quantile_rnaseq = 0.5, we kept the genes whose mean expression or variance is above or equal to the median of the respective distribution. When quantile_rnaseq = 1, we kept no genes. We set quantile_rnaseq to be 0, 0.5 and 0.9. Furthermore, and only in the case where we used the variance as the filtering method, we identified the outliers genes using the $1.5 * \mathrm{IQR}$ rule (3.553 genes), removed them and then applied the quantile_rnaseq filtering. We did not perform the same step when we employed the mean expression, as only ~100 genes were outliers. The above steps happened in the following order: $\log_2$ transformation, outliers removal in the case of the variance filtering method, and gene selection using quantile_rnaseq, either with the mean expression or the variance filtering methods. We used a cross-validation process to identify the best combination of the filtering method and quantile_rnaseq value; see the section Training & Testing protocol below.

Clinical data are available for 409 samples. After consulting our hematology expert, we used the following clinical data. Categorical data included: inferred_ethnicity, consensus_sex, isRelapse, isDenovo, isTransformed, finalFusion, priorMalignancyNonMyeloid, cumulativeChemo, priorMalignancyRadiationTx, priorMDSMoreThanTwoMths, priorMDSMPNMoreThanTwoMths, priorMPNMoreThanTwoMths, ELN2017, specificDxAtAcquisition, specimenGroups, FAB / Blast

Morphology, Karyotype, Other Cytogenetics, Surface Antigens (Immunohistochemical Stains), FLT3-ITD, NPM1, priorMalignancyType, priorMDS, priorMDSMPN, priorMPN, dxAtInclusion, specificDxAtInclusion, dxAtSpecimenAcquisition, specimenType. Numerical data included: ageAtSpecimenAcquisition, %Basophils in PB, %Blasts in BM, %Blasts in PB, %Eosinophils in PB, %Monocytes in PB, Hemoglobin, LDH, Platelet Count, WBC Count, ageAtDiagnosis, timeOfSampleCollectionRelativeToInclusion.

Furthermore, we corrected inconsistent data entries in the following clinical features

- In the "% Blasts in BM" feature, there were three cases where instead of a number, there were ">50" or ">95" data entries. In these cases, we imputed a random number ranging between 51 and 96 to the maximum available number in the data.
- For the "% Blasts in PB" feature, we changed one entry equal to ">90" to a random number between 90.1 and 99.2, two entries equal to "<5" to a random number between 0.1 and 4.9, and three entries equal to "rare" to a random number between 0.1 and 0.9.

### Same input setup

#### Overfitting

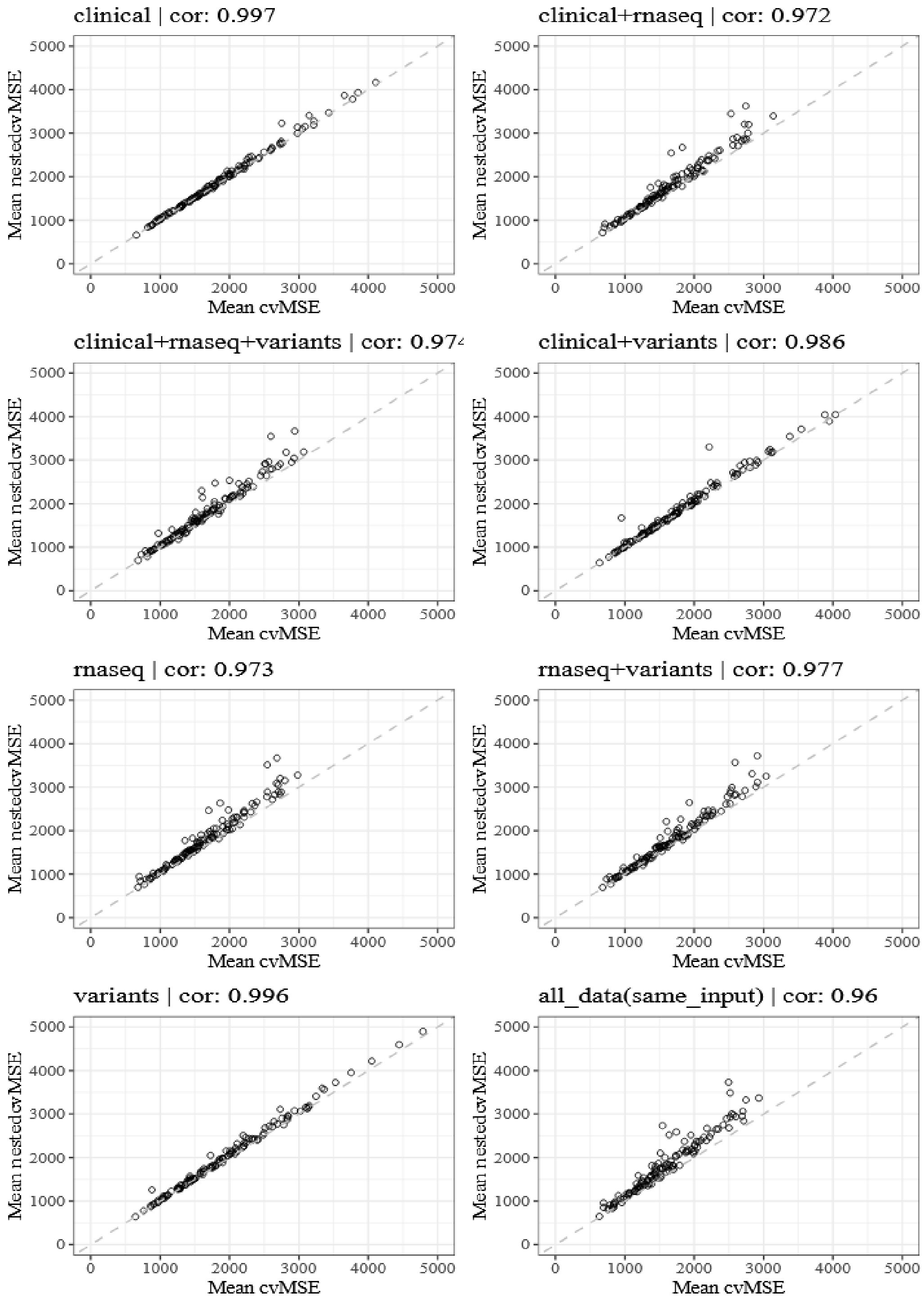


*Supplementary Figure 1. Evaluate overfitting across all drugs, datatypes, and datatypes' combinations.*

#### Statistical tests

| | comparison | pvalue |
|---|---|---|
| 1: | clinical_VS_clinical+rnaseq | 1.961707e-06 |
| 2: | clinical_VS_clinical+rnaseq+variants | 9.017466e-05 |
| 3: | clinical_VS_clinical+variants | 0.7598587 |
| 4: | clinical_VS_rnaseq | 1.389324e-05 |
| 5: | clinical_VS_rnaseq+variants | 0.0002885521 |
| 6: | clinical_VS_variants | 3.73452e-12 |
| 7: | clinical_VS_all_data(same_input) | 0.0001677282 |
| 8: | clinical_VS_all_data(diff_input) | 0.001118324 |
| 9: | clinical+rnaseq_VS_clinical+rnaseq+variants | 0.492325 |
| 10: | clinical+rnaseq_VS_clinical+variants | 5.397458e-06 |
| 11: | clinical+rnaseq_VS_rnaseq | 0.7778793 |
| 12: | clinical+rnaseq_VS_rnaseq+variants | 0.28818 |
| 13: | clinical+rnaseq_VS_variants | 2.7868e-23 |
| 14: | clinical+rnaseq_VS_all_data(same_input) | 0.3326951 |
| 15: | clinical+rnaseq_VS_all_data(diff_input) | 0.1019735 |
| 16: | clinical+rnaseq+variants_VS_clinical+variants | 0.0002670584 |
| 17: | clinical+rnaseq+variants_VS_rnaseq | 0.6998786 |
| 18: | clinical+rnaseq+variants_VS_rnaseq+variants | 0.7737097 |
| 19: | clinical+rnaseq+variants_VS_variants | 1.462212e-19 |
| 20: | clinical+rnaseq+variants_VS_all_data(same_input) | 0.8255547 |
| 21: | clinical+rnaseq+variants_VS_all_data(diff_input) | 0.4055563 |
| 22: | clinical+variants_VS_rnaseq | 3.690592e-05 |
| 23: | clinical+variants_VS_rnaseq+variants | 0.001035505 |
| 24: | clinical+variants_VS_variants | 1.856763e-12 |
| 25: | clinical+variants_VS_all_data(same_input) | 0.0004283569 |
| 26: | clinical+variants_VS_all_data(diff_input) | 0.002547331 |
| 27: | rnaseq_VS_rnaseq+variants | 0.4652969 |
| 28: | rnaseq_VS_variants | 9.966915e-22 |
| 29: | rnaseq_VS_all_data(same_input) | 0.5119677 |
| 30: | rnaseq_VS_all_data(diff_input) | 0.180943 |
| 31: | rnaseq+variants_VS_variants | 6.027028e-19 |
| 32: | rnaseq+variants_VS_all_data(same_input) | 0.951542 |
| 33: | rnaseq+variants_VS_all_data(diff_input) | 0.6313593 |
| 34: | variants_VS_all_data(same_input) | 5.189069e-20 |
| 35: | variants_VS_all_data(diff_input) | 2.833999e-19 |
| 36: | all_data(same_input)_VS_all_data(diff_input) | 0.5225296 |

*Supplementary table 1. Wilcoxon test results, evaluating if the differences of the medians observed in Pearson correlations in Figure 2, achieved across different datatypes or datatypes' combinations, are statistically significant.*

### Performance - Detailed Pearson correlations

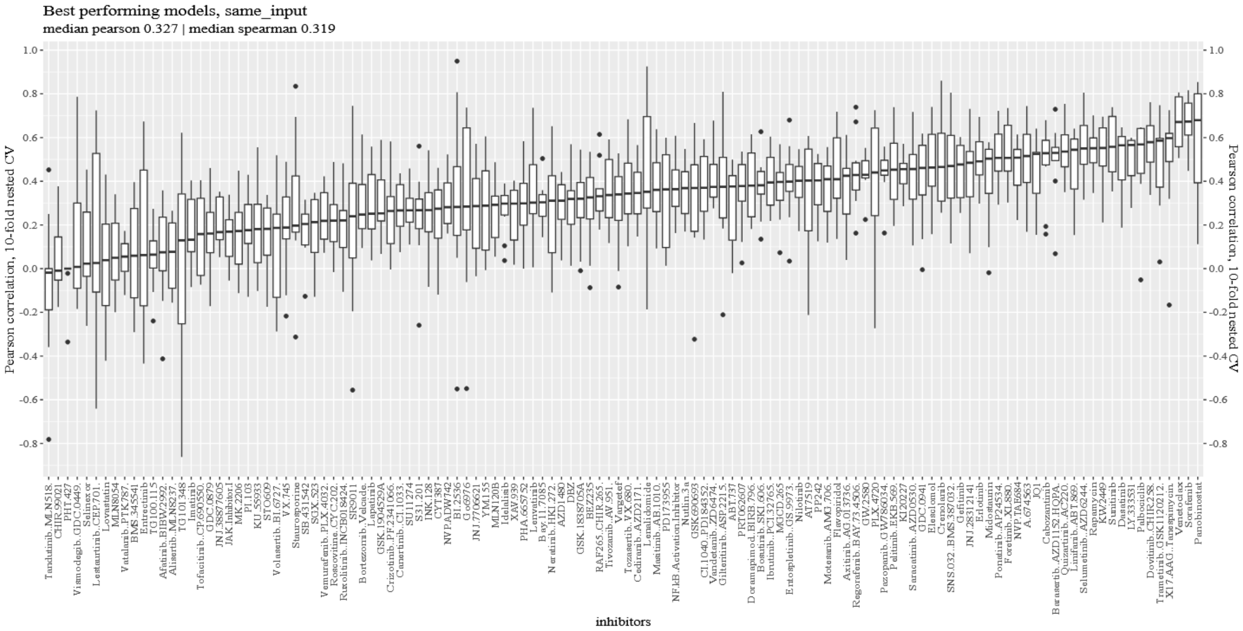


*Supplementary Figure 2. Pearson correlation on the external test set in the 10-fold nested cross-validation in the same_input setting.*

### Different input setup

#### Number of samples

| clinical | clinical+variants | clinical+rnaseq | clinical+rnaseq+variants |
|---|---|---|---|
| Min. : 79.0 | Min. : 74.0 | Min. : 54.0 | Min. : 53.0 |
| 1st Qu.:339.2 | 1st Qu.:307.0 | 1st Qu.:278.2 | 1st Qu.:246.2 |
| Median :347.0 | Median :313.0 | Median :285.5 | Median :253.0 |
| Mean :313.5 | Mean :283.6 | Mean :256.7 | Mean :227.8 |
| 3rd Qu.:352.8 | 3rd Qu.:317.0 | 3rd Qu.:290.0 | 3rd Qu.:256.0 |
| Max. :398.0 | Max. :359.0 | Max. :318.0 | Max. :281.0 |

*Supplementary Table 2. The distribution summary of the available number of samples per inhibitor and datatype.*

### Performance

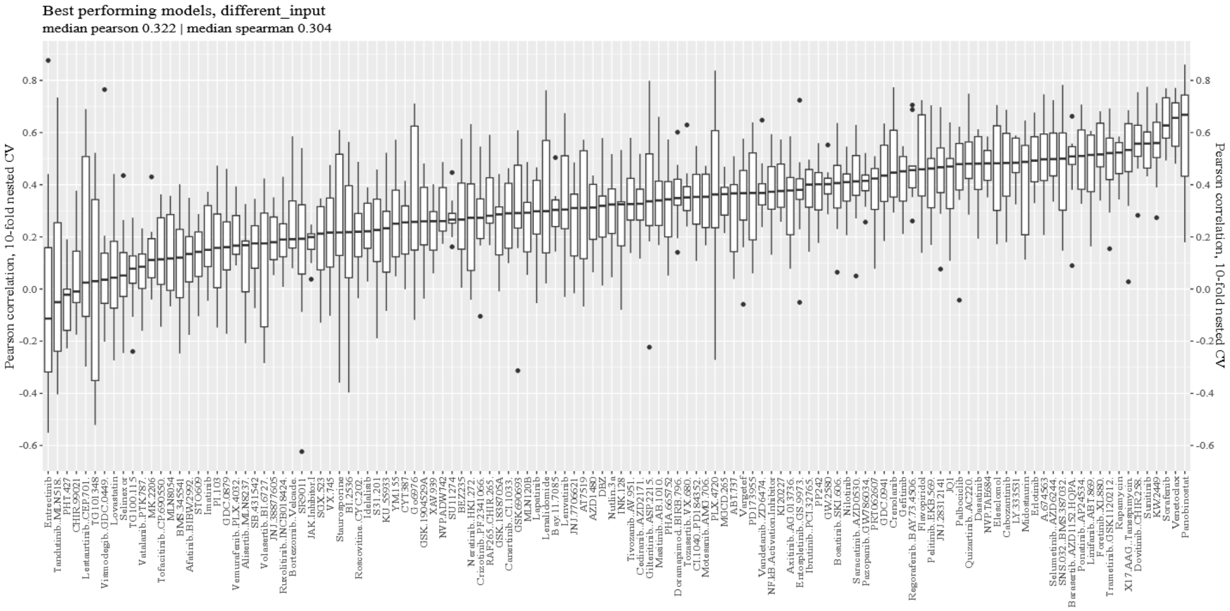


*Supplementary Figure 3. Pearson correlation on the external test set in the 10-fold nested cross-validation in the different_input setting.*

### Models' interpretation

#### Drugs families

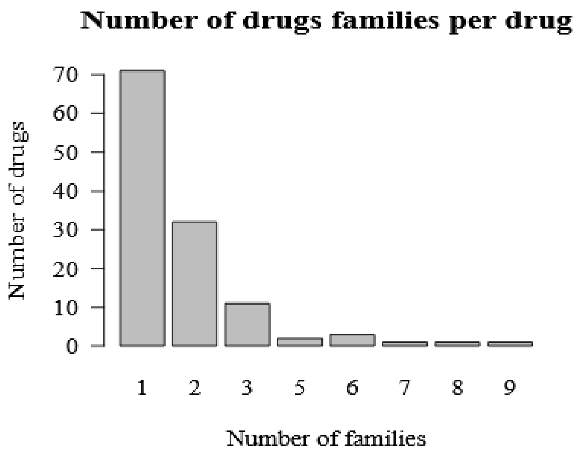


*Supplementary Figure 4. The number of drug families per inhibitor. Seventy-one drugs belong to one drug family, thirty-two drugs belong to two families, etc.*

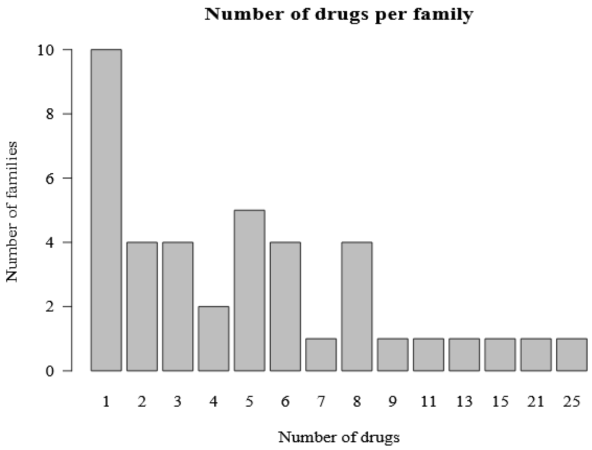


*Supplementary Figure 5. The number of inhibitors per drug family. Ten families have one inhibitor, four families have two inhibitors, etc.*

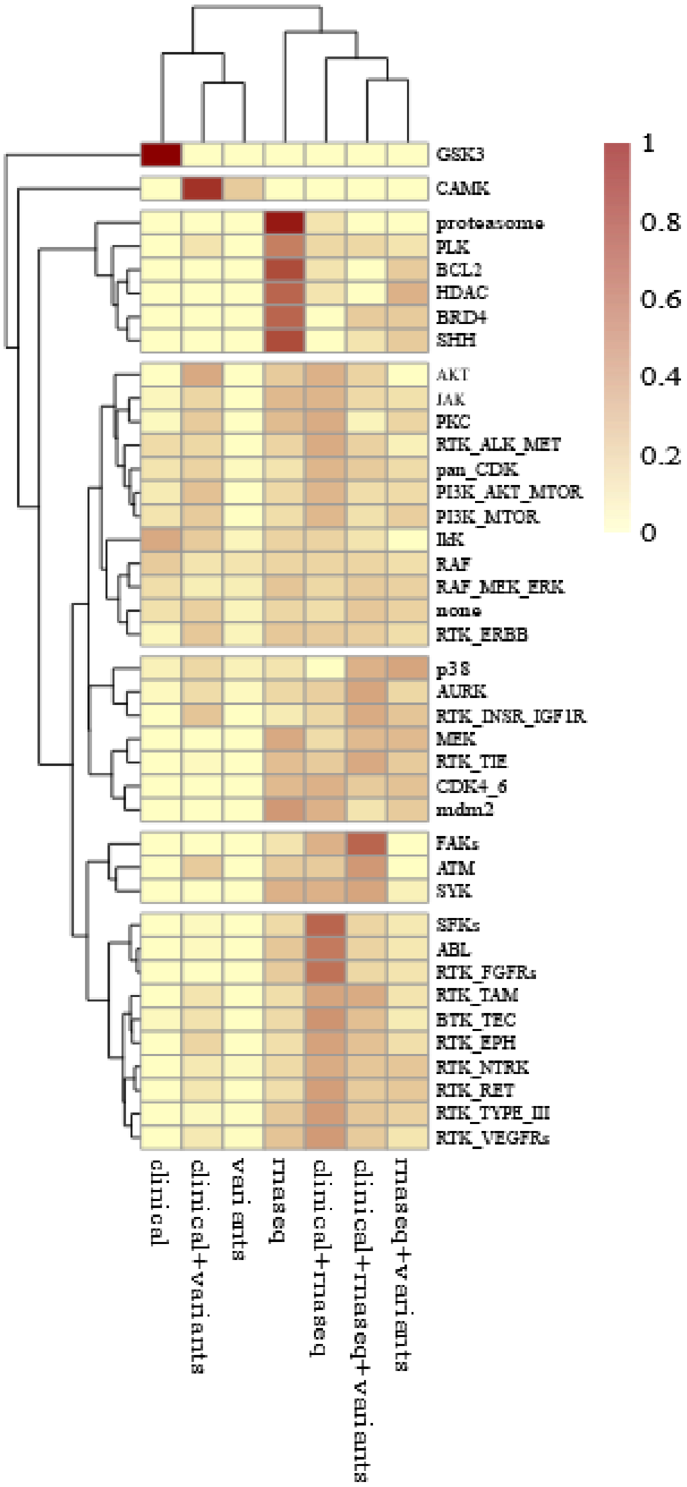


*Supplementary Figure 6. The fraction of the number of times a model selected a datatype or a datatype combination for a drug family during the ten-fold nested cross-validation. Yellow corresponds to a fraction equal to zero, and dark red to a fraction equal to one.*

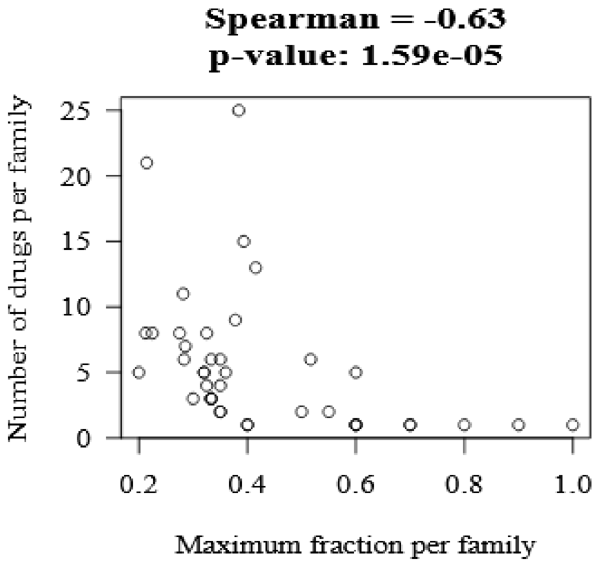


*Supplementary Figure 7. Relationship between the maximum fraction (x-axis) a drug family achieved and the number of its members (y-axis). The fewer the members, the higher the attained fraction, with their Spearman correlation of -0.63 to be highly statistically significant.*

#### Different RNASeq configurations selected for each drug

Here, we focused our analysis on which configurations our models selected when we employed only the RNAseq data for training and testing because they produced the best models when used in isolation. To automate the feature selection and filtering process, we used two different methods, *mean expression* and *variance*, and we optimized the *quantile_rnaseq* parameter. Using *quantile_rnaseq,* we removed a different proportion of the genes incrementally using either their *mean expression* or their *variance* as the filtering method; see section Data in Methods for more details. Our algorithm had the option to filter the data or to keep everything. Filtering helps the performance of our algorithm as it was selected 1061 out of 1220 runs, with *variance* to be chosen 578 times and *mean expression* 483 times, Supplementary Figure 8.

Furthermore, we clustered the drugs based on the times our models selected a filtering method during the nested-cross validation run, Supplementary Figure 9. We observed four main groups. For the drugs in the first group, the *variance* was selected in the majority of the runs. In the second group, the filtering method deviated mainly between the *variance* and the *mean expression*. In the third group, our models selected the *mean expression* in most of the runs. Lastly, in the fourth group, the filtering deviated across all possibilities, *variance, mean expression* and no filtering, with most of the runs selecting no filtering.

Likewise, we evaluated the distribution of the *quantile_rnaseq* parameter corresponding to the proportion of the genes that survived our filtering step, Supplementary Figure 10. In most models, 629 out of 1220, half of the genes were retained (*quantile_rnaseq = 0.5*). Of these 629, 358 used the *mean expression,* and 271 used the *variance*. Interestingly, 315 models out of 1220 used only one-tenth of the genes, with 187 and 125 models using the *variance* and the *mean expression*, respectively. Finally, no filtering or only outliers' removal based on *variance* occurred 159 and 120 times, respectively.

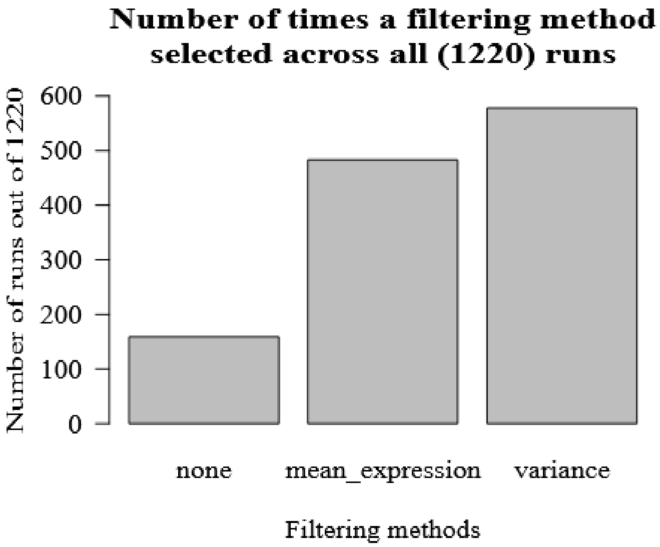


*Supplementary Figure 8. The number of times the models selected a filtering method, mean_expression or variance, across all 1220 runs.*

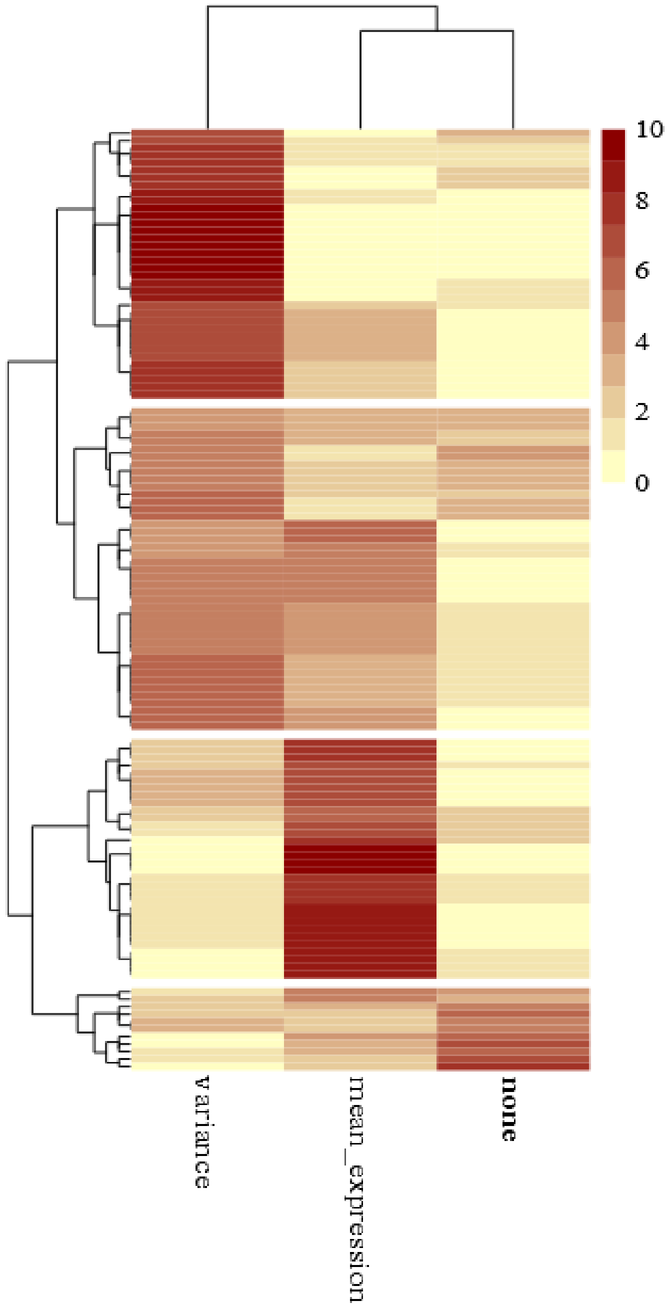


*Supplementary Figure 9. RNAseq filtering methods selection. The number of times a model selected a filtering method for a drug during model building in the ten-fold nested cross-validation. Yellow corresponds to zero times and dark red to ten times. Four main clusters occurred. The first cluster, at the top of the figure, contained drugs that the variance was selected most of the time. The second cluster had drugs that the filtering method deviated between the variance and the mean expression. In the third cluster, the models selected the mean expression more often. In the fourth cluster, the filtering method deviated across all possibilities, and no filtering occurred most of the time.*

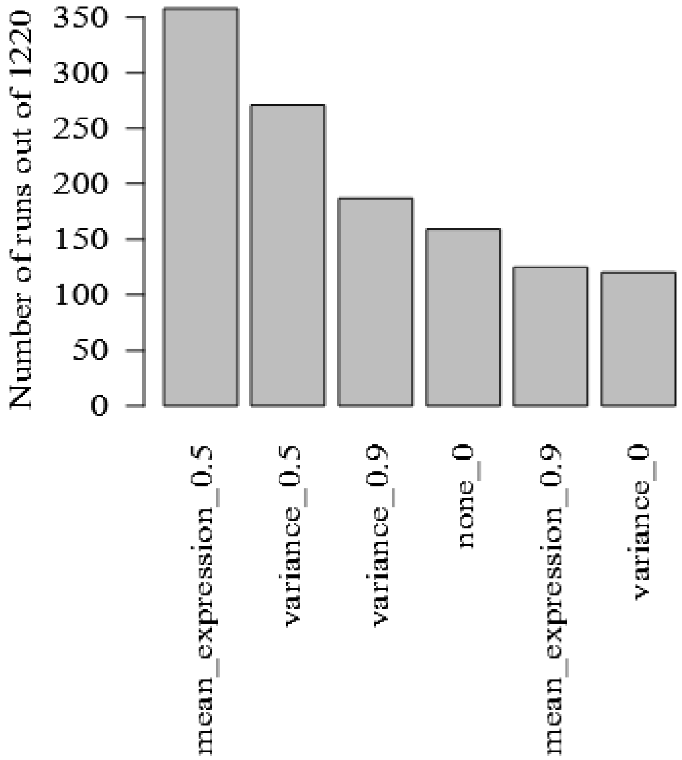


*Supplementary Figure 10. The number of times ElasticNet selected a configuration across all 1220 runs.*

#### Different Whole Exome configurations selected for each drug

Next, we assessed which configurations were selected by our models when we used only whole exome data to train and test our models because it is the primary type of data employed in clinical practice. Our models chose the *tumor_only* filtering option in 650 out of 1220 runs, Supplementary Figure 11. Clustering based on the number of times our models selected the *tumor_only* filtering option, Supplementary Figure 12, resulted in three main clusters. One where the majority of the models selected to use all variants, the second deviated between filtering or not, and the third where our models selected *tumor_only* variants most of the time.

We optimized the *quantile_dnaseq* parameter every time, regardless of the *tumor_only* filtering step. *Quantile_dnaseq* removes a proportion of the variants based on their frequency in the training samples. *Quantile_dnaseq* took three values, 0, where no variants were removed, 0.5 and 0.9 where 50% and 90% of the variants were removed, respectively. Zooming in the cluster, where no *tumor_only* filtering occurred, filtering using *quantile_dnaseq* was beneficial as it was selected 327 times, 133 and 194 times *quanlile_dnaseq* was equal to 0.5 and 0.9, respectively, Supplementary Figure 13. A similar pattern also occurred after *tumor_only* filtering. Our models filtered the variants using *quantile_dnaseq* 331 times, 111 times *quantile_dnaseq* was equal to 0.5 and 220 times to 0.9, Supplementary Figure 14.

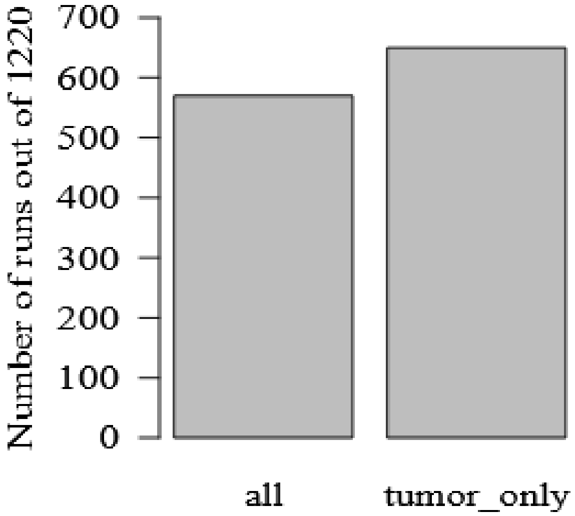


*Supplementary Figure 11. The number of times ElasticNet selected all or the tumor_only variants across all 1220 runs.*

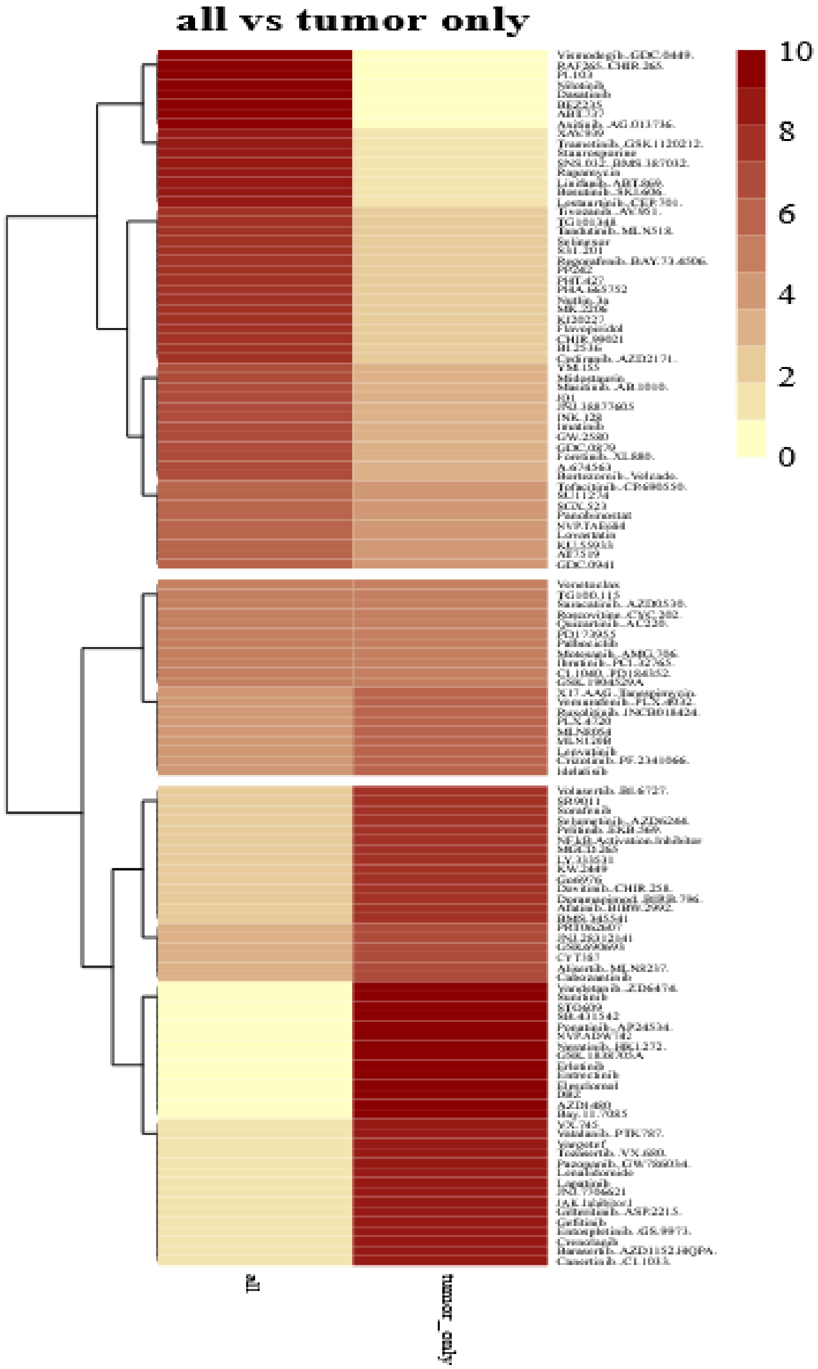


*Supplementary Figure 12. Hierarchical Clustering of the inhibitors based on the number of times the models selected all or the tumor_only variants during model building in the ten-fold nested cross-validation. Yellow corresponds to zero times and dark red to ten times. Three main clusters occurred. Starting from the top, the first cluster contained drugs that used all variants most of the time. The second cluster had drugs that deviated between all and tumor_only variants, while the third cluster included drugs that utilized the tumor_only variants most of the time.*

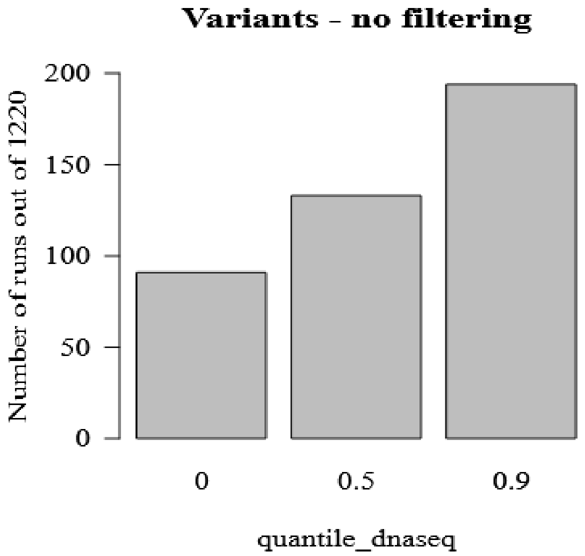


*Supplementary Figure 13. The number of times quantile_dnaseq was equal to 0, 0.5, and 0.9 when focusing on the drugs that belong in the top cluster of Supplementary Figure 11.*

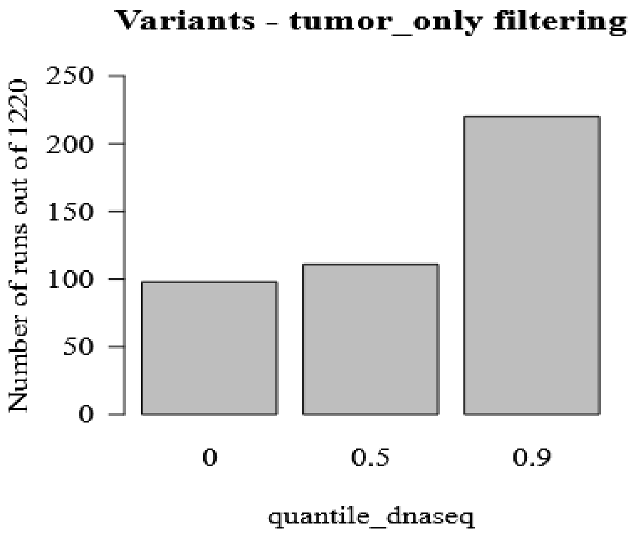


*Supplementary Figure 14. The number of times quantile_dnaseq was equal to 0, 0.5, and 0.9 when focusing on the drugs that belong in the bottom cluster of Supplementary Figure 11.*

### Clinical implications

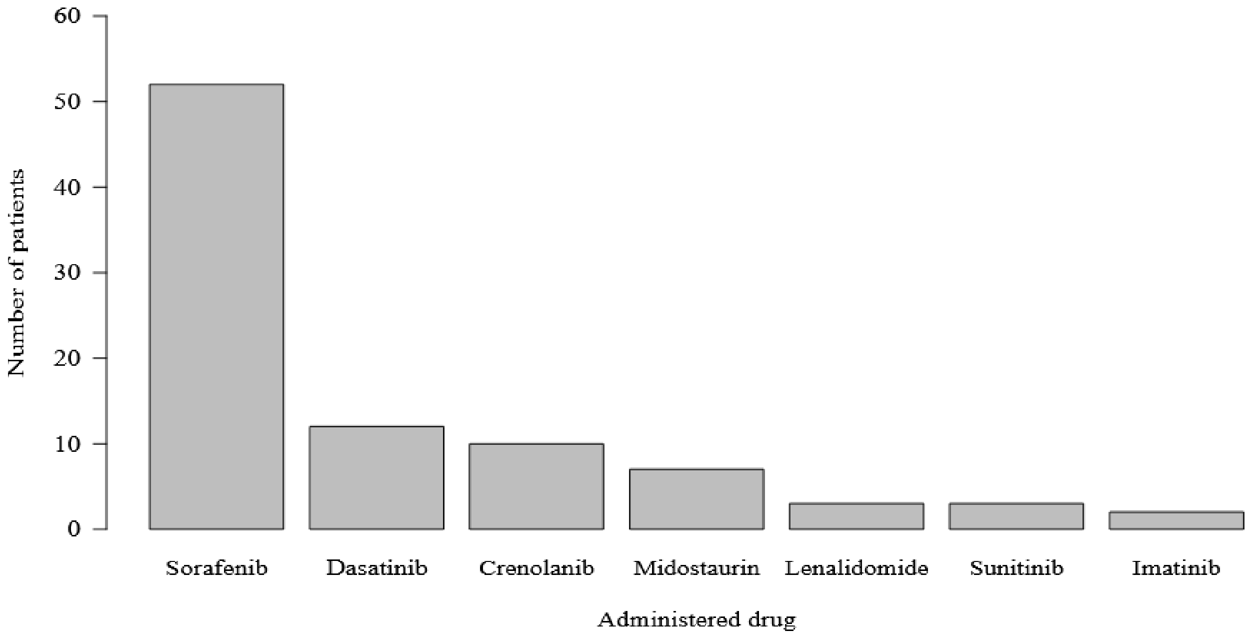


*Supplementary Figure 15. The number of patients that administered a drug and their ex vivo drug response is also available.*

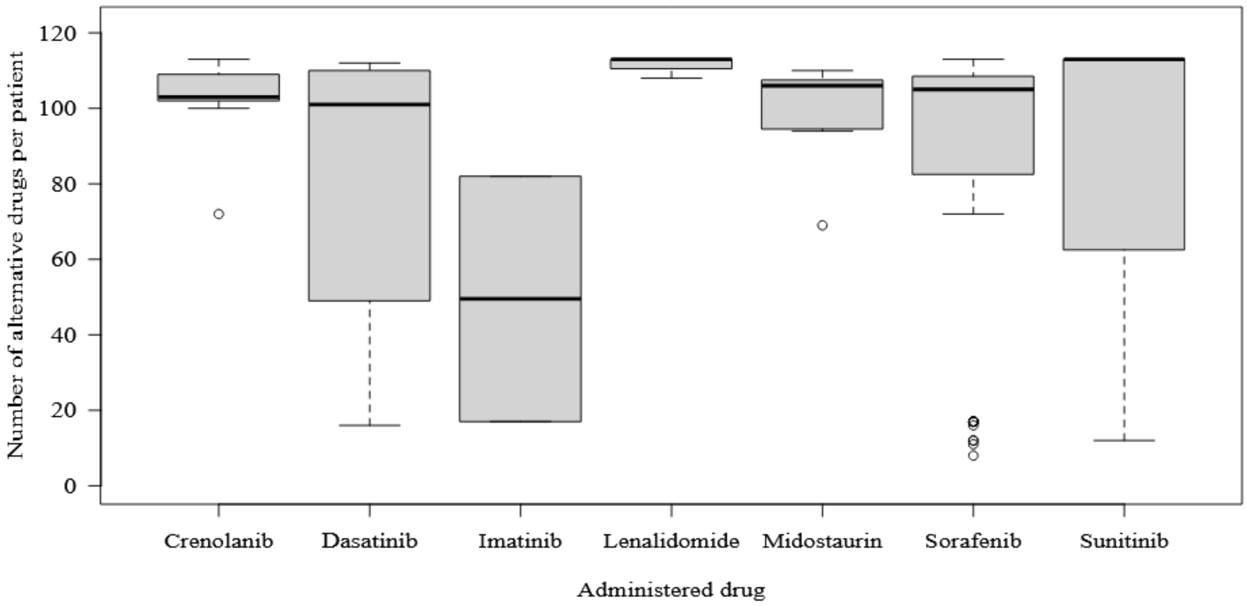


*Supplementary Figure 16. The number of alternative drugs, drugs that have been measured in the same patient as the administered ones. For example, in the case of Crenolanid in most patients, 103 alternative drugs have been measured in the same patient. There is one patient with 72 alternative drugs in the data. The same idea applies to the other boxplots.*

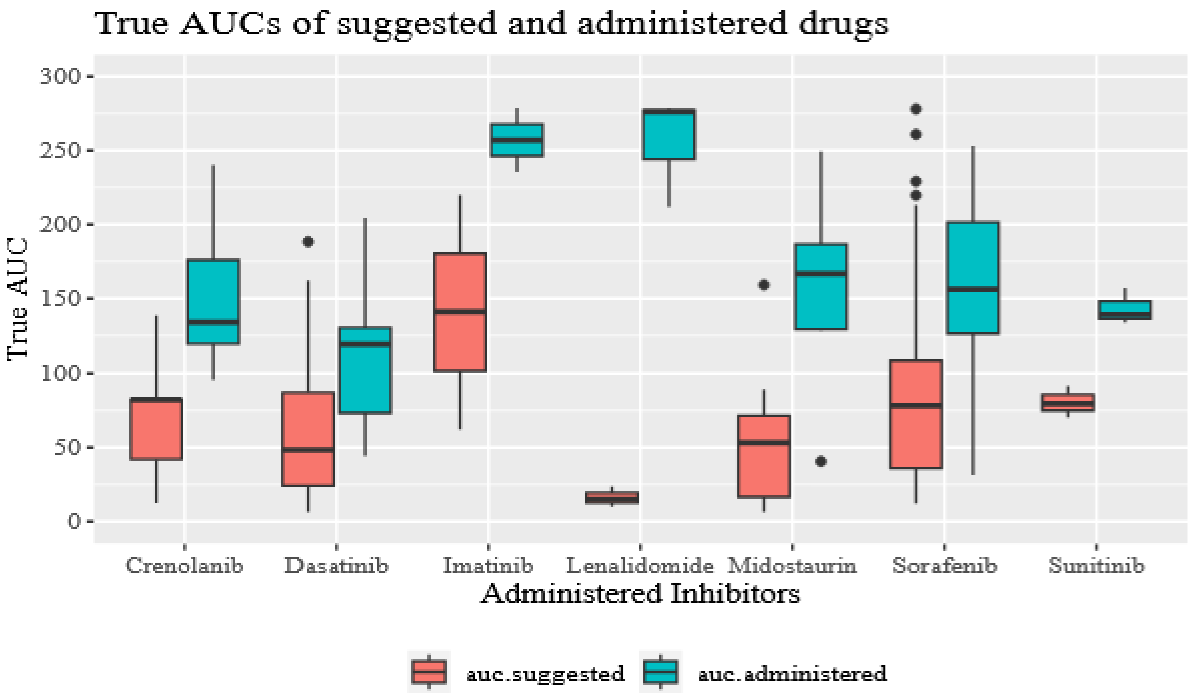


*Supplementary Figure 17. The distribution of the true AUC between the suggested (red) versus the administered (green) drugs.*
